# Machine learning from the CARDAMON trial identifies a carfilzomib-specific mutational response signature

**DOI:** 10.1101/2023.04.08.23288287

**Authors:** Ieuan G Walker, Venetia D’arcy, Garima Khandelwal, Georgina Anderson, Anna Aubareda, William Wilson, Evelyn Fitzsimons, Daria Galas-Filipowicz, Kane Foster, Rakesh Popat, Karthik Ramasamy, Matthew Streetly, Ceri Bygrave, Reuben Benjamin, Ruth M. de Tute, Marquita Camilleri, Selina J Chavda, Gavin Pang, Tushhar Dadaga, Sumaiya Kamora, James Cavenagh, Elizabeth H. Phillips, Laura Clifton-Hadley, Roger G Owen, Javier Herrero, Kwee Yong, Michael A Chapman

## Abstract

Precision medicine holds great promise to improve outcomes in cancer, including haematological malignancies. However, there are few biomarkers that influence choice of chemotherapy in clinical practice. In particular, multiple myeloma requires an individualized approach as there exist several active therapies, but little agreement on how and when they should be used and combined. We have previously shown that a transcriptomic signature can identify specific bortezomib- and lenalidomide-sensitivity. However, gene expression signatures are challenging to implement clinically. We reasoned that signatures based on the presence or absence of gene mutations would be more tractable in the clinical setting, though examples of such signatures are rare. We performed whole exome sequencing as part of the CARDAMON trial, which employed carfilzomib-based therapy. We applied advanced machine learning approaches to discover mutational patterns predictive of treatment outcome. The resulting model accurately predicted progression-free survival (PFS) both in CARDAMON patients and in an external validation set of patients from the CoMMpass study who had received carfilzomib. The signature was specific for carfilzomib therapy and was strongly driven by genes on chromosome 1p36. Importantly, patients predicted to be carfilzomib-sensitive had a longer PFS when treated with carfilzomib/lenalidomide/dexamethasone than with bortezomib/carfilzomib/dexamethasone. However, in those predicted to be carfilzomib-insensitive, the latter therapy may have been capable of eradicating carfilzomib-resistant clones. We propose that the signature can be used to make rational therapeutic decisions and could be incorporated into future clinical trials.

## Introduction

Multiple myeloma has seen an unprecedented increase in active therapies. However, we do not know how these therapies should be best combined, nor in what order they should be employed. Genomic studies have established the inter- and intra-patient heterogeneity of the disease and it is almost certain that the optimal therapy will differ between individual patients and at different lines of therapy. Clinically tractable biomarkers that indicate the sensitivity of individual myeloma subclones to specific drugs would enable therapy combinations to be designed on an individual basis. For example, they could be used to select the best upfront quadruplet regimen in fit patients, or they could inform the most effective doublet or triplet in more frail patients.

Whilst there are plentiful expression or mutational signatures that can identify poor prognosis for specific cancers, including myeloma^1–4^, treatment-specific biomarkers are far less common. Within the latter category, the biomarker is often the direct target of the treatment. Examples are hormone receptors in breast cancer, BRAF V617F for vemurafenib, and BCR-ABL for imatinib and related receptor tyrosine kinase inhibitors. Similarly, a mutation may be part of a synthetic lethal combination, such as BRCA mutations, which confer sensitivity to PARP inhibitors in mutated cancers^5,6^. There are far fewer non-target examples, but these include various indicators of gemtuzumab oligomycin response in acute myeloid leukaemia^7^, and t(11;14) as a marker of venetoclax sensitivity in myeloma^8,9^. This last group of biomarkers were typically discovered in associative studies linking driver mutations known a priori with outcomes in preclinical or clinical trials. Unfortunately, this approach limits their discovery.

We have reasoned that this non-target group of biomarkers represents a large untapped source of drug-specific predictive information, provided we can move beyond simple correlative analyses. Machine learning has found applications in diverse fields and, indeed, we have shown that it can effectively predict drug-specific responses in myeloma. Our seven-gene expression signature not only predicted bortezomib- and lenalidomide-specific outcome, but illustrated a reciprocity of response between these drugs, implying that any combination effect seen in clinical trials was an effect of targeting of different clones, rather than through synergistic activity^10^. Whilst transcriptional prognostic signatures have been employed in patient care^11–13^, they are generally challenging to translate to the clinic^14^, largely because of normalization issues. Mutational signatures are much simpler to interpret.

The CARDAMON (Carfilzomib/Cyclophosphamide/Dexamethasone with Maintenance Carfilzomib in Untreated Transplant-eligible Patients with Symptomatic MM to Evaluate the Benefit of Upfront ASCT) trial was a phase II trial exploring upfront carfilzomib/cyclophosphamide/dexamethasone (KCD), followed by randomization to an autologous high-dose stem cell transplant or KCD consolidation, followed by carfilzomib maintenance (**Supplementary Fig. 1**)^15^. As CARDAMON was a clinical trial employing carfilzomib as the single novel anti-myeloma agent, we reasoned that it could be employed to train a machine learning algorithm to learn carfilzomib responsiveness. We employed a highly successful ensemble machine learning algorithm to predict for progression-free survival (PFS), then independently validated this signature in The Multiple Myeloma Research Foundation-sponsored Relating Clinical Outcomes in Multiple Myeloma to Personal Assessment of Genetic Profile (CoMMpass) trial and further showed that our model could influence clinical decision making.

## Results

### Whole exome sequencing in CARDAMON produced a representative genomic dataset

We performed whole exome sequencing of somatic and germline samples of 148 patients from CARDAMON. Clinical and laboratory characteristics of these patients are shown in **Supplementary Table 1**. Mean coverage was 115X (somatic) and 35X (germline). All but three samples passed quality control. Copy number profiles, non-synonymous single nucleotide variants, pattern of variants, and variants per sample were all consistent with previous genomic studies in myeloma^16–20^; **Supplementary Fig. 2**). Having established that our dataset was of a high quality and representative, we proceeded to search for carfilzomib-specific biomarkers.

### Development of a machine learning model to predict PFS after carfilzomib treatment

The impact of single gene mutations on treatment outcome have been explored previously in a homogeneously treated patient group, as part of the Myeloma XI trial^2^. TP53, ATM, and ATR mutations were associated with prognosis, almost certainly a treatment-independent result, but ZNFHX4, IRF4, and EGR1 all predicted response to (predominantly) immunomodulatory (IMiD) therapy^21^. However, absence of an appropriate external validation group precluded independent testing of these. Following these results, we examined the impact of known driver mutations on KCD response in CARDAMON, but we saw no correlation between these and PFS (data not shown).

We therefore proceeded to establish a machine learning model using XGBoost ^22^, an ensemble tree machine learning algorithm, which has dominated recent Kaggle competitions (https://github.com/dmlc/xgboost/tree/master/demo#machine-learning-challenge-winning-solutions). We trained a model to regress PFS on the presence or absence of gene mutations (whether clonal or sub-clonal) in patients receiving KCD. Our best model in validation contained 17 trees, each with six nodes (a node representing presence or absence of a mutation), encompassing a total of 61 genes. We investigated whether any of the genes selected by XGBoost had any biological relevance. Interestingly, there was a significant enrichment within a specific chromosomal location; 21 of the 61 genes utilised in the model exist on chromosome 1p36 (p=3.11×10^−18^) which has previously been described as a poor risk marker in myeloma^24^.

### The model predicts carfilzomib-specific prognosis in an external dataset

The CoMMpass dataset^25,26^ is a major resource for independent validation of genomic results in myeloma. We tested our model on the 274 patients who had received carfilzomib-based combinations first-line in CoMMpass. Because of the heterogeneity of this patient group compared to the CARDAMON training cohort, we did not expect the model’s predictions to be accurate in an absolute sense (e.g. a patient with a predicted PFS of two years would not necessarily be expected to progress at two years exactly). Rather, we anticipated that we could use predicted PFS in a relative manner (e.g. a patient predicted to progress at two years should progress after a patient predicted to progress at one year). We thus separated the patients into prognostic groups. We first split the patients into approximately two equal-sized groups (based on *predicted* PFS of greater or less than two years), a predicted carfilzomib-sensitive and a predicted carfilzomib-insensitive group, and compared their observed survival. The median PFS was not reached (95% confidence interval [CI] 59.0 months – not reached) in patients predicted to be carfilzomib-sensitive and 36.0 months (95% CI 24.9-70.6) in patients predicted to be carfilzomib-insensitive (hazard ratio [HR] 2.39; 95% CI 1.32 – 4.32; p=0.004; **Fig. 1A**). We also performed a comparison of PFS in three risk groups based on the model’s predictions (upper quartile, middle two quartiles, and lower quartile of predicted PFS). Median PFS was not reached (95% CI not obtainable) in the carfilzomib-good risk group; median PFS was not reached (95% CI 58.9 – not reached) in the carfilzomib-intermediate risk group (HR 2.31; 95% CI 1.03 – 17.25; p = 6.0×10^−4^; **Fig. 1B**); and was 35.5 months (95% CI 23.3 – 62.2) in the carfilzomib-poor risk group (HR 6.73; 95% CI 1.87 – 49.9; p=8.0×10^−4^; **Fig. 1B**).

**Figure 1.**
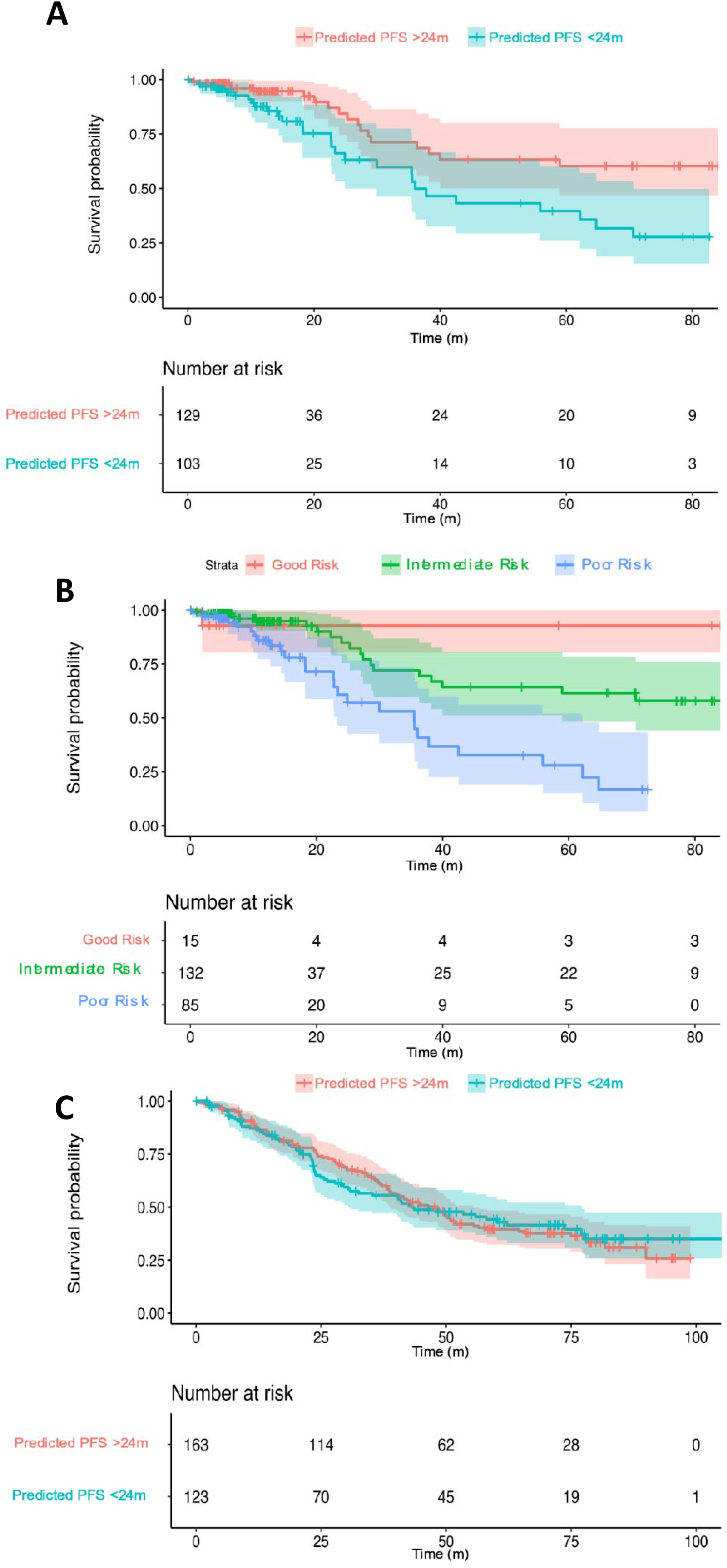
External validation of our model demonstrates that it specifically predicts carfilzomib-sensitivity. (A) Kaplan-Meier of progression-free survival (PFS) of patients predicted by the model to be carfilzomib-sensitive (red) or carfilzomib-insensitive (cyan) and who were treated with carfilzomib-based therapy in CoMMpass. (B) Kaplan-Meier of PFS of the same patients in (A), stratified by the model into three groups of carfilzomib-sensitivity: low-risk (green), medium-risk (blue), high-risk (red). (C) Kaplan-Meier of PFS of patients predicted by the model to be carfilzomib-sensitive (red) or carfilzomib-insensitive (cyan) and who were treated with bortezomib-based therapy in CoMMpass.

We wanted to check whether our model was acting as a generic prognostic signature, i.e. whether it would predict for response regardless of therapy. We therefore predicted carfilzomib-sensitivity using our model in CoMMpass patients treated with bortezomib-based therapy, reasoning that, if the model were carfilzomib-specific, PFS should be similar in the two groups. Indeed, this was the case. The median PFS was 48.3 months (95% CI 38.7 – 56.5) in patients predicted to be carfilzomib-sensitive and 43.3 months (95% CI 30.8 – 77.1) in patients predicted to be carfilzomib-insensitive (HR 1.01; 95% CI 0.73 – 1.38; p = 0.96; **Fig. 1C**). Taken together, these data suggest that our model predicts outcome in a carfilzomib-specific manner.

### The model can inform clinical decisions

Carfilzomib has been shown to be superior to bortezomib in the second-line setting^27^. Therefore, the outcome of the ENDURANCE trial^28^, in which the PFS of patients treated with bortezomib/lenalidomide/dexamethasone (VRD) or carfilzomib/lenalidomide/dexamethasone (KRD) was not significantly different, was surprising. We wondered if rational therapy selection might have altered the outcome of that trial. We hypothesized that in CoMMpass, carfilzomib-sensitive patients treated with KRD would have superior PFS to carfilzomib-sensitive patients receiving VRD. Conversely, we predicted that VRD would be superior to KRD in carfilzomib-insensitive patients. But first, we wanted to ensure that the data in CoMMpass mirrored those of ENDURANCE by comparing all-comers treated with VRD with those treated with KRD. Indeed, PFS was not significantly different between the two groups. There were 421 patients treated with VRD and 176 patients treated with KRD. The median PFS was 50.4 months (95% CI 36.3 – not reached) in patients treated with KRD and 43.8 months (95% CI 40.3 – 48.4) in patients treated with VRD (HR 0.80; 95% CI 0.58 – 1.09; p=0.16; **Fig. 2A**).

**Figure 2.**
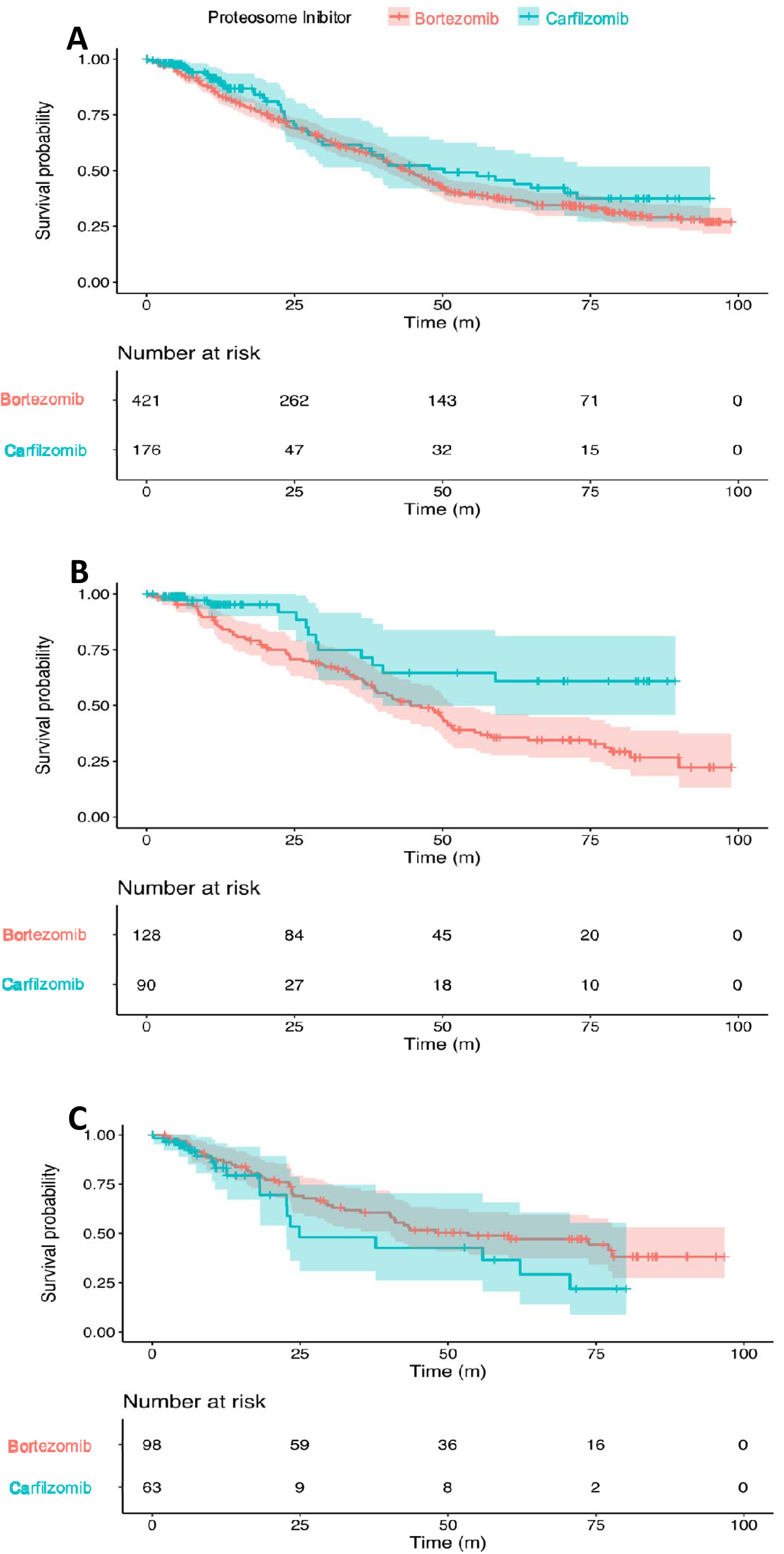
XGBoost model accurately predicts response to Carfilzomib in patients predicted to respond. (A) Kaplan-Meier plot of progression-free survival (PFS) of patients who received carfilzomib/lenalidomde/dexamethasone (KRD) first-line (cyan) versus patients who received bortezomib/lenalidomide/dexamethasone (VRD) first-line (red) in CoMMpass. (B) Kaplan-Meier plot of PFS of patients who were predicted to be carfilzomib-sensitive by our model, split by which proteosome inhibitor they received, either KRD first-line (cyan) or VRD first-line (red) in CoMMpass. (C) Kaplan-Meier plot of PFS of patients who were predicted to be carfilzomib-insensitive by our model, split by which proteosome inhibitor they received, either KRD first-line (cyan) or VRD first-line (red) in CoMMpass.

We then predicted carfilzomib-sensitivity for all patients in the CoMMpass database treated first-line with KRD or VRD who had associated whole exome sequencing data. Of the VRD patients, 128 were predicted to be carfilzomib-sensitive and 98 were predicted to be carfilzomib-insensitive. Of the KRD patients, 90 were predicted to be carfilzomib-sensitive and 63 were predicted to be carfilzomib-insensitive. Within these two groups, for patients predicted to be carfilzomib-sensitive, the median PFS was not reached (95% CI 39.9 – not reached) in patients treated with KRD and 44.6 months (95% CI 38.6 - 51.9) in patients treated with VRD (HR 0.43; 95% CI 0.24 – 0.77; p= 0.0045; **Fig. 2B**). For patients predicted to be carfilzomib-insensitive, the median PFS was 53.4 months (95% CI 40.2 – not reached) in patients treated with VRD and 24.9 months (95% CI 22.7 – not reached) in patients treated with KRD (HR 1.52; 95% CI 0.88 – 2.64; p= 0.14; **Fig. 2C**). Thus, for patients predicted to be carfilzomib-sensitive, there appears to be a clear survival advantage with KRD treatment over VRD treatment. For patients predicted to be carfilzomib-insensitive, the PFS was higher with VRD than KRD, although this difference was not significant. The ENDURANCE trial demonstrated that VRD had fewer cardiac, renal or pulmonary related adverse events than VRD. On the other hand, the trial also showed that 17% of VRd patients vs 10% of KRd patients discontinued therapy due to adverse reactions. This shows that it is critically important to deliver the correct drug to the correct patient to maximize the benefit:risk ratio, which our model now allows.

### Carfilzomib-insensitive sub-clones can be effectively treated with other drugs

To further explore whether our model could usefully inform clinical decisions, we explored predicted carfilzomib-sensitivity at a sub-clonal level. Of 142 patients with more than one temporally spaced whole exome samples in CoMMpass, we identified 66 patients who had carfilzomib-insensitive clones at diagnosis. Patients who received carfilzomib (N = 9), either diagnosis, saw their carfilzomib-insensitive sub-clone expand at relapse (**Figs. 3A** and **3B**). However, interestingly, there were 11 patients whose carfilzomib-resistant clone disappeared below the limit of detection following treatment (**Fig. 4**). 8/11 of these patients had received lenalidomide as part of their treatment, and a further two underwent high dose melphalan autografts. This provides further evidence that. If used in clinical practice, the model could be used to influence clinical decisions.

**Figure 3.**
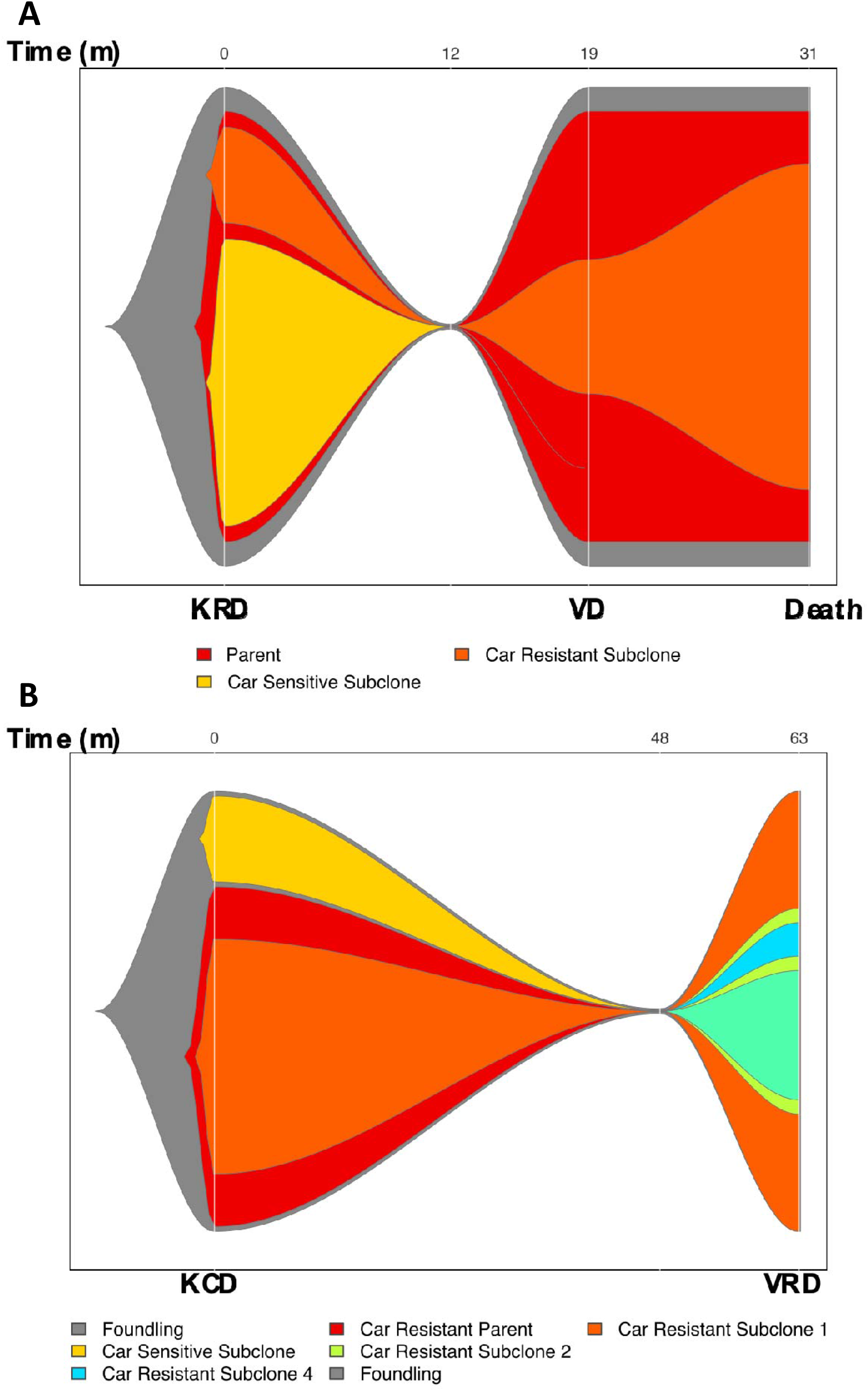
Carfilzomib resistant subclones expand when treated with Carfilzomib. (A) & (B) Fish plots of patients who have subclones predicted to be carfilzomib-insensitive at diagnosis. Both were treated with carfilzomib-based regimes first-line. Y-axis denotes proportion of clonal cancer frequency, and X-axis time.

**Figure 4.**
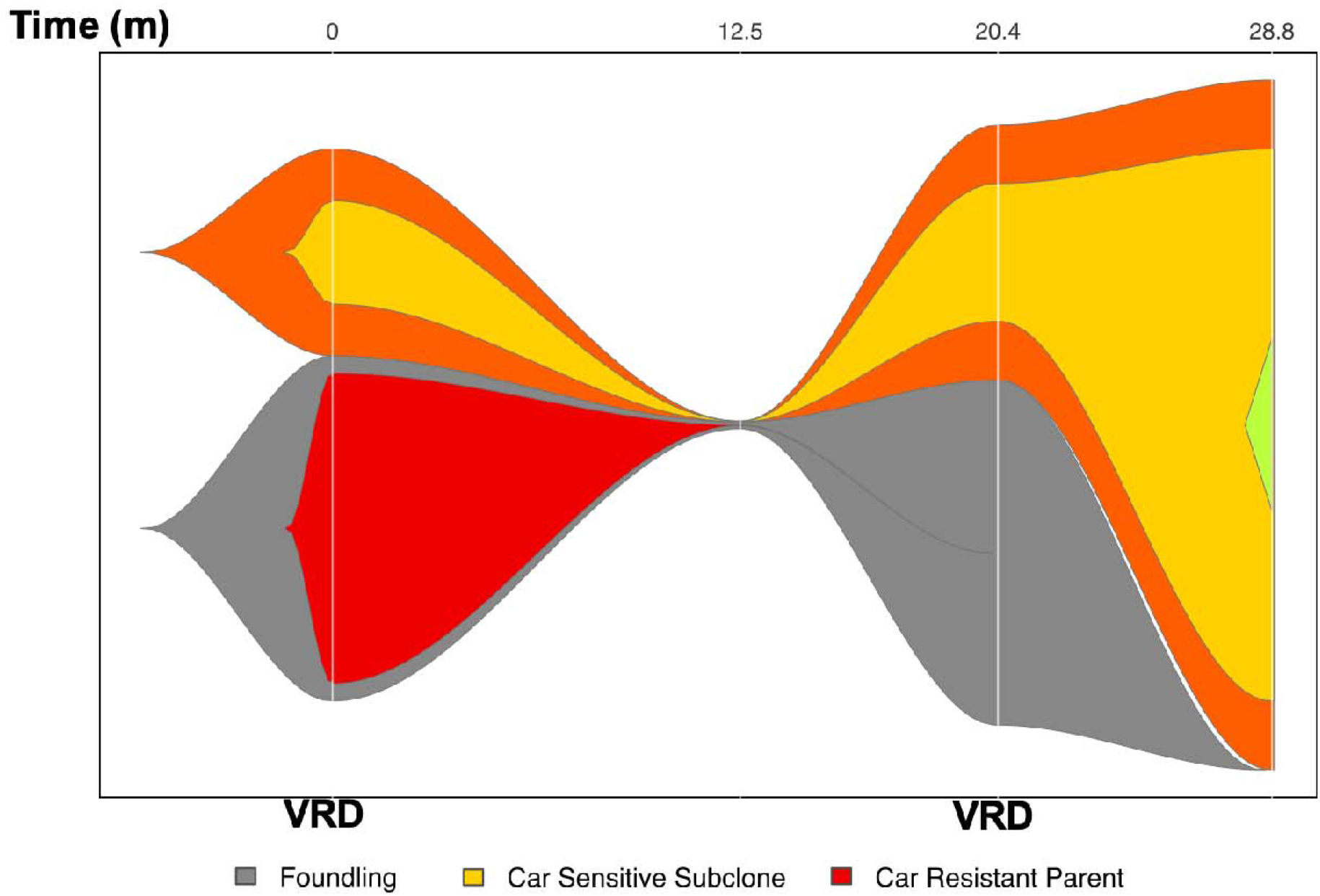
Carfilzomib resistant subclones respond to alternative therapies. Fish plot of patient with carfilzomib-resistant clone at diagnosis treated with bortezomib/lenalidomide/dexamethasone (VRD). Y-axis denotes proportion of clonal cancer frequency, and X-axis time.

## Discussion

It is now considered that, for patients who are fit enough, upfront quadruplet therapy with an IMiD, a proteasome inhibitor, an anti-CD38 monoclonal antibody, and high-dose steroid is the treatment of choice, either in combination with a high-dose melphalan autograft^29–31^ or without^32^. However, it is not clear what is the best combination of drugs, nor is it likely that the same drug combination will benefit all patients. Furthermore, whilst phase 3 randomized control trials remain the sine qua non of evidence-based medicine, they are extremely expensive, cannot cover all comparisons, and are currently inadequate to address precision approaches.

The ENDURANCE trial showed no difference in efficacy between VRD and KRD, even though carfilzomib-based therapies have demonstrated better efficacy than bortezomib-based therapies second-line^27^. Using a mutation-based signature, which is much easier to implement in “real-time” than transcriptomic signatures, we show here that we can distinguish between patients who are likely to benefit from carfilzomib-based therapy and those who are not. The message of the ENDURANCE trial was that VRD was preferable to KRD because the former was not inferior in terms of PFS but was associated with lower rates of severe and a subset of cardiac-renal-and-pulmonary adverse events. This paper suggests that, had the patients received VRD or KRD according to our model (VRD for those carfilzomib-insensitive, KRD for those carfilzomib-sensitive), outcomes would have been significantly better. To be clear, we would not necessarily anticipate any difference in PFS between the rationally treated VRD or KRD patients within this hypothetical trial if treated accordingly to our model. Instead, we would anticipate that the PFS of all patients within the hypothetical trial would have been significantly higher than those reported in the original trial. As exome data were collected as part of ENDURANCE, it should be possible to answer this question by comparing the outcome of patients receiving the “correct” predicted therapy versus those receiving the “incorrect” predicted therapy.

A major challenge will be designing clinical trials that show the added value of precision medicine models prospectively. We not only have to show that receiving the correct predicted treatment outperforms the incorrect treatment, but that the act of stratifying patients by some rational model is, by itself, beneficial. One possibility would be to perform add-on studies to existing large phase 3 trials. For example, consider the existing RADAR trial^33^. In this trial, patients could be prospectively evaluated with a targeted 61 gene exome library that enables our model to be employed. Then, a sub-group of patients deemed carfilzomib-sensitive could be randomized to receive KRD or to continue with the standard VRD. A second sub-group of patients deemed carfilzomib-insensitive could be randomized in the same way. This would provide all the data to make the necessary comparisons, including whether model stratification itself was beneficial.

Machine learning is becoming increasingly influential in all aspects of life. Within medicine, there are clinical areas, such as interpretation of radiographs^34–36^ and pathology slides^37–39^, that are very close to being realized by artificial intelligence. For these fields, extremely high accuracy is required to supplant experienced radiologists and histopathologists. This, in turn, requires very large numbers of training samples. In contrast, for cancer therapy, because we are trying to improve on current paradigms, where treatment choices are based on blanket applications of a treatment regimen shown to be beneficial in one arm of a clinical trial, without stratification – but realistically, also based on whether a regimen is licensed and funded and whether the patient is fit enough to receive it – we have shown that rational treatment predictions that perform better than chance can significantly improve survival. Thus, whilst we do not have the training numbers or accuracy of machine learning in histopathology or radiology, our approach has huge potential for improving outcomes of cancer treatment without the need for new drugs. Prospective testing of this approach is urgent and critical.

## Methods

### Whole exome sequencing (WES) library preparation

All patients underwent WES of germline and myeloma samples following CD138+ bead selection. All DNA samples were prepared using Agilent SureSelectXT Low Input Kit/Human All Exon V6 baits with UTRs: 5190-8882, and sequenced on an Illumina 3000 HiSeq instrument (Illumina, San Diego, CA) according to the manufacturer’s recommendations. Alignment was performed against Hg38 using the Burrow’s Wheeler-MEM^40^.

### Mutation calling

Somatic mutations were called using Mutect2^41^ according to best practices (https://software.broadinstitute.org/gatk/best-practices/) to generate VCF files.

### RNAseq library preparation

Patients underwent library prep using mRNA (Poly A+) enrichment using the Kapa mRNA HyperPrep Kit. Sequencing was done on Illumina 3000 HiSeq instrument (Illumina, San Diego, CA) according to the manufacturer’s recommendations. Adapter sequences were removed using Cutadapt and quality control was performed using FastQC. Alignment was done using STAR aligner to the Hg38 genome. Gene quantification and differential expression was performed using SAMTools feature counts and DESeq2.

### Machine Learning and Statistical analysis

All downstream analysis was performed using R (R Core Team (2021). R: A language and environment for statistical computing. R Foundation for Statistical Computing, Vienna, Austria. URL https://www.R-project.org/), version 4.1.2. Mutations were then expanded to gene level (presence of a mutation within any exon of the gene). The machine learning algorithm, XGBoost^22^, was then used to predict accelerated failure time^23^. The following parameters were derived by initial 80:20 validation experiments: eta = 0.1, tree method = hist, max_depth = 6, eval_metrics = aft-nlog and standard error, nrounds = 17.

Survival curves were generated using the Kaplan-Meier estimate and multivariate analysis was performed using Cox’s proportional hazard model.

### Subclonal analysis

Clonal structure of patients was defined using Sciclone^42^. Foundling and subclones were assigned by their relative clonal cancer frequency. Fish plots to show sub-clonal structure were generated using Fishplots^43^.

### Data and Code Availability

Data from the COMMpass trial was accessed using the following url: https://research.themmrf.org using version IA20.

All code used for the preparation of this manuscript is publicly available at the following GitHub URL: https://github.com/ieuangw/CARDAMON_ML. VCF files will be uploaded to GEO upon peer-reviewed publication. A summary table of all mutations in cancer samples is included in supplemental table 1.

## Data Availability

All data produced in the present study are available upon reasonable request to the authors. Data and code will be made publicly available on acceptance in a peer reviewed journal

## Acknowlegements

IGW is supported by the Kay Kendall Leukaemia Fund (KKL1442) also received support from the UK Myeloma Society (UKMS), Vd’A has received support from the UKMS. MAC is supported by the Medical Research Council Toxicology Unit (MC_UU_00025/10). KY and RP are supported by the National Institute for Health Research University College London Hospitals Biomedical Research Centre. The CARDAMON trial was funded by Amgen and endorsed by Cancer Research UK (C9203/A17750). The CARDAMON trial was developed within the National Cancer Research Institute Myeloma Working Group and is part of the UK Myeloma Research Alliance portfolio. We gratefully acknowledge the patients and carers who participated in this trial.

## Supplementary Figure legends

**Supplemental Figure 1.**
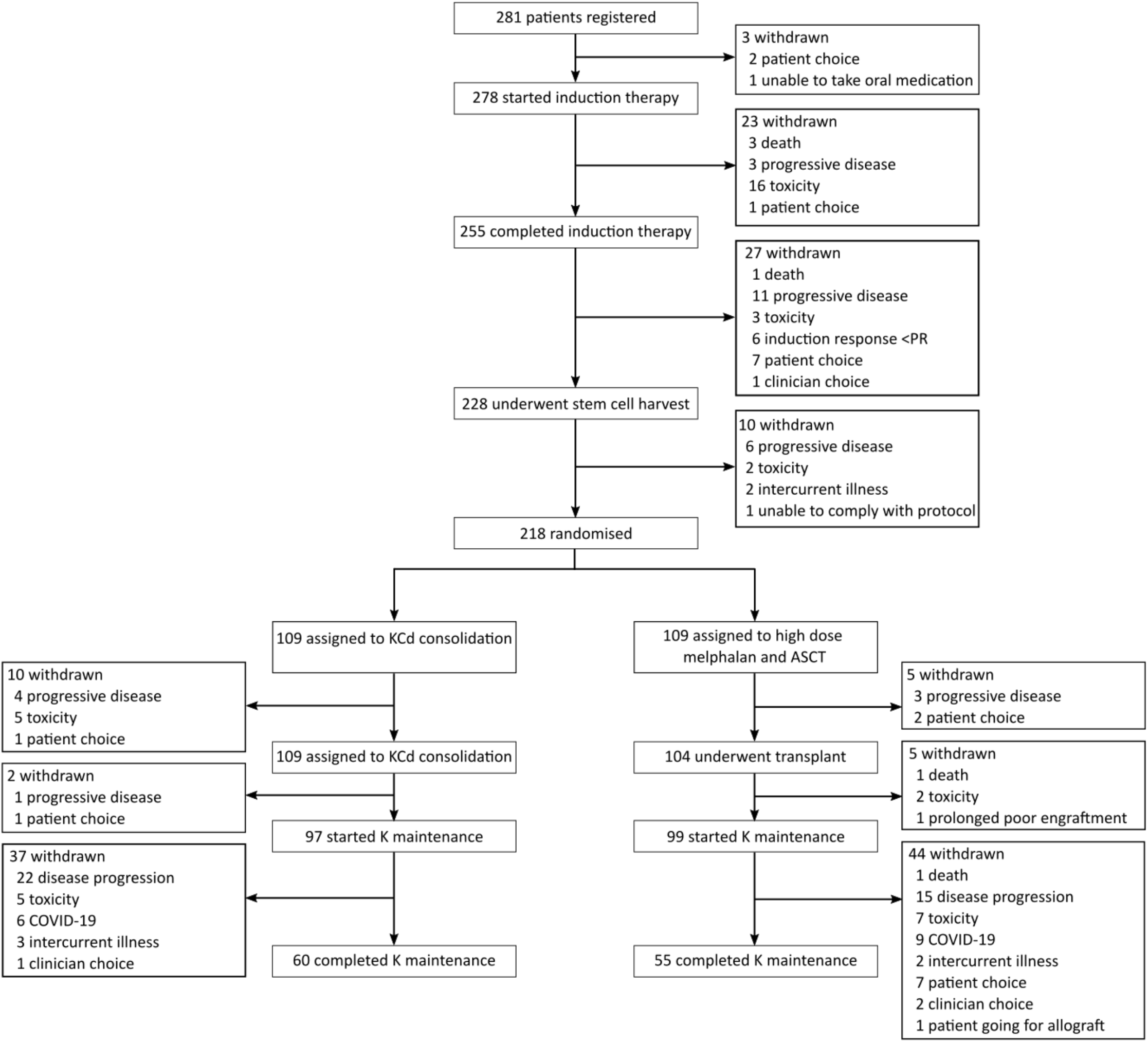
CONSORT diagram of the CARDAMON Trial. All numbers are intention to treat.

**Supplemental figure 2.**
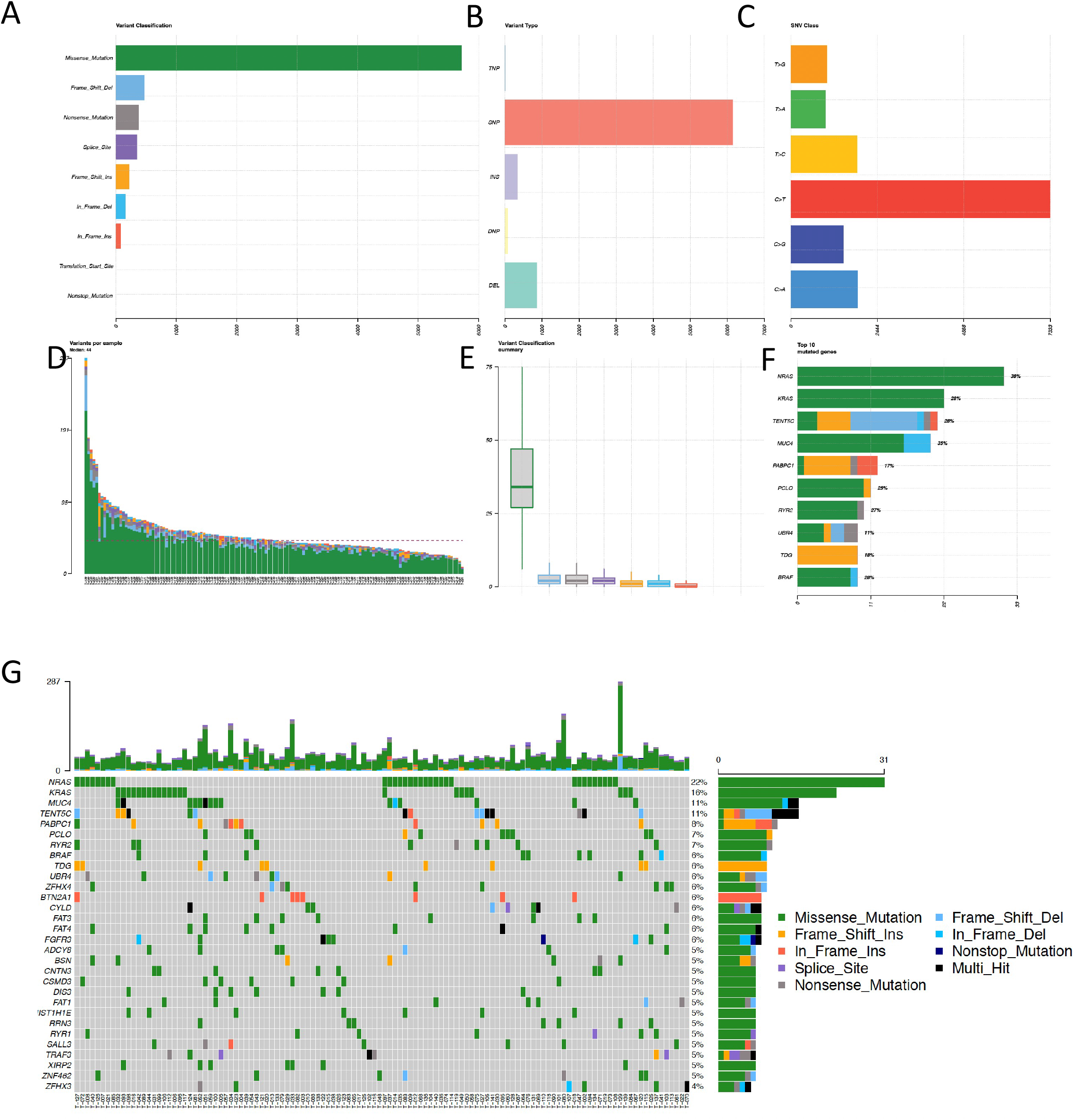
CARDAMON whole exome sequencing data are high-quality and representative. (A) Variant classification summaries. Top left – number of reads annotating each mutation type call called by MUTECT2. Top middle – frequency of variant type called by MUTECT2. Top right – base mutation change frequency. Bottom left – number of mutations detected per patient. Bottom middle – variant classification summary. Bottom right – the top 10 genes mutated in patients in the CARDAMON trial. (B) Heatmap showing presence or absence of top 30 most frequently mutated genes. Rows represent genes, columns represent patients. Grey = no mutation detected for that patient.

