## Supplemental Figure legends for "Machine learning from the CARDAMON trial identifies a carfilzomib-specific mutational response signature"

**Supplemental figure 1. Consort diagram of the CARDAMON Trial.** A consort diagram of the CARDAMON trial, all numbers are as intention to treat. Data correct at time of publication of Primary end point of the trial.

**Supplemental figure 2. Whole Exome Sequencing summary of the CARDAMON Trial. A)** Number of reads annotating each mutation type call called by MUTECT2. **B)** Frequency of variant type called by MUTECT2 **C)** Base mutation change frequency **D)** Number of mutations detected per patient X axis, colour denotes type of mutation called by MUTECT2. **E)** Variant classification summary **F)** the top 10 genes mutated in patients in the CARDAMON trial. **G)** Heatmap showing presence or absence of top 30 most frequently mutated genes. Rows genes, columns are patients. Grey = no mutation detected for that patient.
